# Amplicon based MinION sequencing of SARS-CoV-2 and metagenomic characterisation of nasopharyngeal swabs from patients with COVID-19

**DOI:** 10.1101/2020.03.05.20032011

**Authors:** Shona C. Moore, Rebekah Penrice-Randal, Muhannad Alruwaili, Xiaofeng Dong, Steven T. Pullan, Daniel P. Carter, Kevin Bewley, Qin Zhao, Yani Sun, Catherine Hartley, En-min Zhou, Tom Solomon, Michael B. J. Beadsworth, James Cruise, Debby Bogaert, Derrick W. Crook, David A. Matthews, Andrew D. Davidson, Zana Mahmood, Waleed Aljabr, Julian Druce, Richard T. Vipond, Lisa F. P. Ng, Laurent Renia, Peter J. M. Openshaw, J. Kenneth Baillie, Miles W. Carroll, Malcolm G. Semple, Lance Turtle, Julian A. Hiscox

## Abstract

COVID-19 is a complex disease phenotype where the underlying microbiome could influence morbidity and mortality. Amplicon and metagenomic MinION based sequencing was used to rapidly (within 8 hours) identify SARS-CoV-2 and assess the microbiome in nasopharyngeal swabs obtained from patients with COVID-19 by the ISARIC 4C consortium.

## Introduction

Mechanisms of life-threatening disease in COVID-19 including factors associated with morbidity and mortality. Middle East respiratory syndrome (MERS) can provide important parallels: one of the risk factors associated with severe disease and/or a fatal outcome is the presence of other infections [1, 2]. Other infections may therefore exacerbate COVID-19. A case study in China identified the presence of other microorganisms in patients with COVID-19 and included specific nucleic acid based detection for six common respiratory virus and the use of culture [3]. No viruses were identified but the study identified *Acinetobacter baumannii, Klebsiella pneumoniae*, and *Aspergillus flavus* in a single patient and several cases of fungal infection were diagnosed including *Candida albicans* and *Candida glabrata*. The ability to rapidly identify both the primary pathogen and other infections that may be present early in disease may provide opportunities for targeted intervention in patients suspected or confirmed with COVID-19. The advantage of both laboratory [4] and field-based sequencing approaches [5] was illustrated in the West African Ebola virus outbreak in characterising viral infection and also providing an assessment of the contribution of potential co-infections to outcome [6]. Rapid sequencing of SARS-CoV-2 itself provides utility in two main areas. First, specific amplicon-based sequencing of SARS-CoV-2 allows for potential contact tracing, molecular epidemiology and studies of viral evolution. Second, the use of metagenomic approaches like SISPA provides a check on sequence divergence for amplicon-based approaches. This is particularly important for SARS-CoV-2 as the virus could undergo recombination with other human coronaviruses and mutation and this may also affect both vaccine and antiviral efficacy.

## Methods

### Ethics and clinical information

Patients used in this study gave informed consent and were recruited under the International Severe Acute Respiratory and emerging Infection Consortium (ISARIC) Clinical Characterisation Protocol CCP (https://isaric.net/ccp), reviewed and approved by the national research ethics service, Oxford (13/SC/0149). Sequencing approaches were validated on samples from two patients, Patient 1 who was asymptomatic and Patient 2 who experienced a 4-day flu-like illness. Samples were timed from the day of recruitment.

### RNA extraction and preparation

Nasopharyngeal swabs collected from patients with COVID-19 were placed in a viral transport medium and RNA isolated using QIAamp viral RNA mini kit (Qiagen) by spin-column procedure according to the manufacturer’s instructions. Total RNA was purified from SARS-CoV-2 infected Vero cells using the Qiagen RNA minikit following AVL inactivation. Infection of Vero cells was conducted at Containment Level 3 at Public Health England, Porton Down. RNA samples were treated with Turbo DNase (Invitrogen).

### Primer design

The SARS-CoV-2 reference sequence from NCBI (NC_045512.2) was aligned to 16 SARS-CoV-2 sequences published on GISAID. Primer binding sites were chosen according to defined conserved regions after alignment. Primers, available by request, were chosen that sequentially amplified roughly 1000 bp with an ∼200 bp overlapping region.

### RT-PCR

SuperScript IV was used to generate single strand cDNA using random hexamers. The primer sets were used to generate 30 amplicons from the cDNA. The reaction conditions were; denaturation at 98°C for 30 sec followed by 35 cycles of 10 sec denaturation at 98°C, 30 sec annealing at 66°C, and then 50 sec of extension at 72°C. A final extension step was done for 2 min at 72°C.

### SISPA

For round A, RNA was reverse-transcribed with SuperScript IV Reverse Transcriptase (Invitrogen) using Sol-PrimerA (5′-GTTTCCCACTGGAGGATA-N9-3′), followed by second-strand DNA synthesis with Sequenase DNA polymerase 2.0 (Invitrogen). For the round B reaction, Round A-labeled cDNA was added to the Q5-high-fidelity 2xmaster mix (NEB) per sample with Sol-PrimerB (5′-GTTTCCCACTGGAGGATA-3′).

### Library preparation for MinION sequencing

PCR products were purified using AMPure XP beads (Beckman Coulter [A63880]). NEBNext FFPE DNA Repair Mix (M6630) and NEBNext End repair/dA-tailing module (E7546) reagents were added to the PCR mix. Following end-repair of the amplicons, ligation buffer, NEBNext Quick T4 ligase (E6056) and Adapter mix (AMX [ONT]) were added. Amplicons with ligated adapters were then quantified on the qubit fluorometer before loading onto the flow cell. One flow cell was used for each sample with no multiplex.

### EPI2ME (WIMP)

Fast5s generated by the MinION sequencer were base called into fastqs by Guppy. Fastq files were uploaded to Oxford Nanopore Technology’s (ONT’s) cloud-based pipeline EPI2ME (WIMP rev. 3.2.2) workflow to retrieve information about the identity of the nucleic acid in the clinical samples.

## Results

To identify and sequence SARS-CoV-2 two complementary techniques were used. First, an amplicon-based system and the second, a metagenomic approach, that can also be used to assess the background microbiome. For the amplicon-based system, a series of primers were designed such that the SARS-CoV-2 genome could be amplified in appropriately 1000 base paired sequential fragments, with an approximately 200 base pair overlap to allow sequence assembly from the amplicon data (Fig. 1A). The primers were selected on the basis of conserved regions in the SARS-CoV-2 genome based upon an initial deposition of 17 genomes. The rationale to generate short amplicons was also to allow the selection of primer pairs that could amplify longer segments should one set of primers fail on a particular sample. To test whether the primers could generate amplicons, RNA was purified from Vero cells that had been infected with an isolate of SARS-CoV-2 (MT007544.1 GenBank). This RNA was used as a template for cDNA synthesis followed by PCR using the conserved primers. This generated 30 amplicons covering the SARS-CoV-2 genomes and these were sequenced using MinION (Fig. 1B). The amplicon-based approach was then evaluated on nasopharyngeal swabs collected from two patients with COVID-19. Three samples were used - Patient 1 – Days 1 and 3 and Patient 2 – Day 1. Amplicons were generated and sequenced using the MinION with reads mapping to SARS-CoV-2 (Fig. 1C). The data indicated that potentially some amplicons were potentially more abundant than others, but nevertheless resulted in identification and sequencing of the viral genome.

**Fig. 1.**
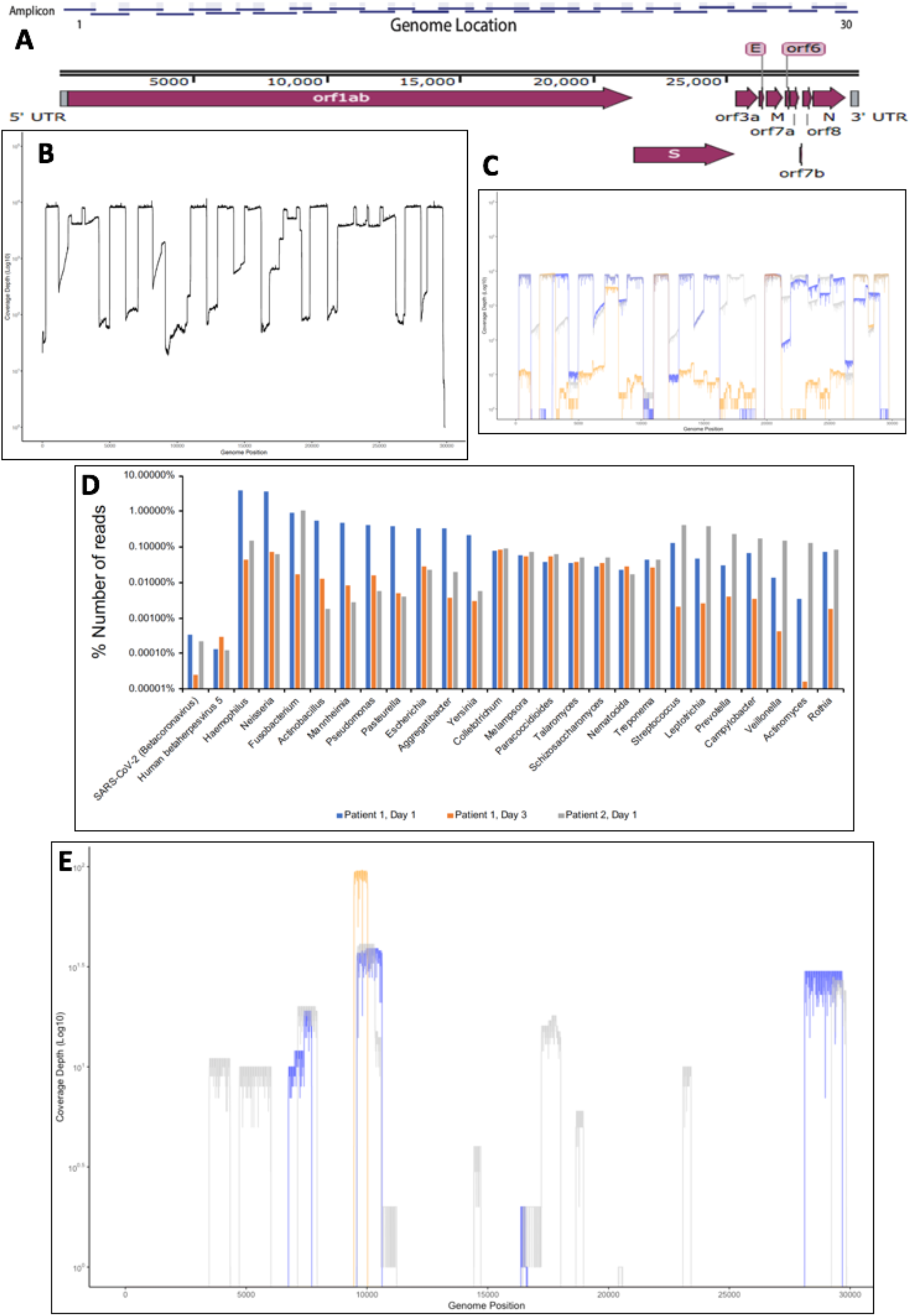
(A) Schematic diagram of the SARS-CoV-2 genome with the position of the amplicons illustrated above. (B). Validation and sequence read depth analysis of amplicons generated from RNA isolated from Vero cells infected with SARS-CoV-2. (C). Sequence read depth analysis of amplicons generated from RNA isolated from patients with COVID-19. Blue is Patient 1, Day 1, orange is Patient 1, Day 3, and grey is Patient 2, Day 1. (D). Proportion of reads for each patient mapping to SARS-CoV-2 and other identified nucleic acids for bacteria, virus and fungi using a metagenomic approach. (E). Location of mapped sequences on the SARS-CoV-2 genome using the metagenomic approach, Blue is Patient 1, Day 1, orange is Patient 1, Day 3, and grey is Patient 2, Day 1.

To assess the respiratory microbiome, and to provide an alternative approach to identify SARS-CoV-2 in a clinical sample, a metagenomic approach was used. This made no prior assumptions as to what nucleic acid was present in the RNA extracted from the oropharyngeal swabs. Here, amplification by sequence-independent single primer amplification (SISPA) was used. This had previously been used to identify dengue, chikungunya, influenza and Lassa fever viruses in clinical samples [7-9]. To provide an internal control, samples were spiked with nucleic acid from a plasmid expressing the VP35 RNA from Ebola virus (EBOV) [10]. Samples were not bar coded and each sample was processed on a single flow cell. Sequence reads were mapped to retrieve information about the identity of the nucleic acid in the clinical samples. The total reads classified were 8,698,559, 9,890,327 and 5,849,966 for Patient 1 – Day 1, Patient 1 – Day 3 and Patient 2 – Day 1, respectively. Unsurprisingly, the majority of reads mapped to the human genome with 6,833,956, 9,661,370 and 5,422,187, for Patient 1 – Day 1, Patient 1 – Day 3 and Patient 2 – Day 1, respectively (data not shown). Nucleic acid mapping to the spike-in was identified in each sample and was correctly identified by the mapping software, providing confidence in the approach (data not shown). Nucleic acid mapping to bacteria was also identified and was used to identify at both the genus level (Fig. 1D) and species level as well. For example, in in Patient 2 – Day 1, 58,645 reads were mapped to *Fusobacterium periodonticum*. Reads mapping to viruses could also be identified and these included SARS-CoV-2 and human betaherpes virus 5 (human cytomegalovirus) (Fig. 1D). Coverage of the SARS-CoV-2 genome in the three samples was not uniform (Fig. 1E) and unsurprisingly the read depth was much lower than with the amplicon-based approach (Fig. 1C).

## Discussion

This work demonstrates that amplicon-based sequencing is feasible for the study of the SARS-CoV-2 genome in samples from patients. Although not formally timed, we estimate that using this approach coupled to MinION based sequencing, then genomic information can be obtained within ten hours. One of the limitations is working at higher containment. We would note that our base calling used Epi2ME and artefacts are often observed by Kraken. BLAST analysis provides greater accuracy but takes longer. Sequencing of viral genomes during outbreaks provides much needed information in terms of viral adaptation [4] and informs molecular epidemiological studies [11]. Due to recombination in coronaviruses (e.g. [12, 13]) current diagnostics may not remain fit for purpose and therefore metagenomic approaches provide independent verification of the presence of viral genomes as well information on the underlying microbiome – which may contribute to severe disease in COVID-19. One limitation of the metagenomic approach is the limit of detection and in this study, not all of the SARS-CoV-2 genome was sequenced using the SISPA approach. For diagnostic purposes, RT-qPCR generally is more sensitive, provided that the primer binding sites remain conserved in the pathogen being tested. In this case, RT-qPCR diagnostic reagents can be revaluated based upon using sequencing as sentinel for these events. Determining the background microbiome in near real time can inform potential treatment strategies in the event specific co-infections are identified. Future efforts will quantify the limits of detection using genome sequencing by these approaches.

## Data Availability

Data can be obtained from the corresponding author by request

## Author contributions

Conceptualisation: M.W.C., M.G.S., J.K.B., P.J.M.O., W.A., L.T. and J.A.H. Methodology: S.M., R.P.-R., L.T., M.A., X.D., D.W.C., Z.M., D.A.M., R.V., W.A., S.T.P. and A.D.D. Software: R.P.-R., X.D. and D.A.M. Validation: S.C.M., R.P.-R. and M.A. Formal analysis and investigation: S.C.M., R.P.-R., M.A., X.D., S.T.P., D.B., D.W.C., D.P.C., J.K.B., Q.Z., Y.S., C.H. and J.D. Resources: S.T.P., D.P.C., J.K.B., M.B.J.B., J.C., L.T., L.F.P.N., L.R., D.A.M., A.D.D. and J.D. Data Curation: R.P.-R. and X.D. Writing - Original Draft: S.C.M. R.P.-R., M.A., L.T. and J.A.H. Writing - Review & Editing: All authors. Visualization: R.P.-R., X.D. and M.A. Supervision: J.A.H., L.T., M.W.C., E.-M.Z. and W.A. Funding acquisition: J.A.H., M.W.C., W.A., T.S., L.T., J.K.B. P.J.M.O. and M.G.S.

